# SARS-CoV-2 shedding, infectivity and evolution in an immunocompromised adult patient

**DOI:** 10.1101/2021.06.11.21257717

**Authors:** Maria Cassia Mendes-Correa, Fábio Ghilardi, Matias Chiarastelli Salomão, Lucy S. Villas-Boas, Anderson Vicente de Paula, Tania Regina Tozetto-Mendoza, Wilton Freire, Flavia C. Sales, Camila M. Romano, Ingra M. Claro, Leandro Menezes de Souza, Jessica F. Ramos, Heuder Gustavo Oliveira Paiao, Roberta Shcoinik Szor, Esper Georges Kallas, Ester C. Sabino, Steven S. Witkin, Nuno R. Faria

**Affiliations:** Instituto de Medicina Tropical, Faculdade de Medicina da Universidade de São Paulo, São Paulo, Brazil; Departamento de Molestias Infecciosas e Parasitarias da Faculdade de Medicina da Universidade de São Paulo; Hospital 9 de Julho; Hospital das Clínicas da Faculdade de Medicina da Universidade de Sao Paulo; Centro de Oncologia Hospital Sírio Libanês; Weill Cornell Medicine, USA; Department of Infectious Disease Epidemiology, Imperial College London, UK; Department of Zoology, University of Oxford, UK

**Author notes:** **Corresponding Author** Maria Cassia Mendes-Correa, Faculdade de Medicina da Universidade de Sao Paulo-Laboratorio de Investigacao Medica em Virologia (LIM52)-Instituto de Medicina Tropical de Sao Paulo. These authors contributed equally (Maria Cassia Mendes-Correa, Fábio Ghilardi).

**Keywords:** SARS-CoV-2, cell culture, evolution, immunodeficiency, shedding

## Abstract

This report describes a persistent SARS-CoV-2 infection of at least 218 days in a male, in his 40’s who had undergone a prior autologous hematopoietic stem cell transplant due to a diffuse large B-cell lymphoma. He did not manifest a humoral immune response to the virus. Whole-genome sequencing and viral cultures confirmed a continual infection with a replication-positive virus that had undergone genetic variation for at least 196 days following symptom onset.

## Introduction

Chronic infection of SARS-CoV-2 in immunocompromised individuals has been associated with within-host viral evolution and immune escape. However, data regarding the replication potential, shedding and virus diversity in long-term persistent infections in immunocompromised individuals remains scarce [1-3]. No specific infection control guidelines exist regarding immunocompromised individuals. The present report describes an immunocompromised patient with persistent shedding of SARS-CoV-2 for a long period after primary infection, and whose replication-competent virus accrued an extensive number of mutations consistent with intra-host viral genomic evolution.

### Case presentation

We describe the case of a persistent SARS-CoV-2 infection in a male, in his 40’s with a past medical history of autologous hematopoietic stem cell transplantation (HSCT) to treat a diffuse large B-cell lymphoma. In 2020, six months after HSCT while on maintenance therapy with acyclovir, sulfamethoxazole, trimethoprim, glucocorticoids and cyclo-phosphamide, the patient became aware of a fever (37.8° C), myalgia and headache. He then sought medical assistance and underwent a clinical evaluation. Vital signs and pulmonary auscultation were normal. On the same day he underwent blood and swab tests and was positive for SARS-CoV-2 by nasopharyngeal reverse transcription polymerase chain reaction (RT-PCR). A chest computer tomography (CT) scan revealed interstitial pneumonia restricted to basal lung areas. He was discharged with oral antibiotics (amoxicillin/clavulanate plus azithromycin). A worsening of his symptoms led him back to the hospital where he was admitted as an inpatient. At this time his oxygen saturation (SpO2) was 87%. During this first hospitalization he received corticosteroids (prednisone) 0.5 mg/kg and a five-day course of ceftriaxone and azithromycin.

His clinical status and laboratory findings improved and he was finally discharged from hospital at the end of the month. The patient had regular follow-ups and did not report any new signs or symptoms. He kept using prednisone at gradually reduced doses. A few days latter, he noticed new signs of dyspnea with fatigue and chest pain and also complained about having a fever although this was not measured. This clinical worsening lead to a 2nd hospitalization for three months .This prolonged stay was due to a relapse in his interstitial pneumopathy. There was a rapidly increased oxygen requirement associated with a worsening clinical condition, lymphopenia and a low monocyte count. He also developed several bacterial infections related to this prolonged hospitalization.

Due to his severe condition and persistence in being RT-PCR-positive for SARS-CoV-2, he was treated with plasma from individuals who had recovered from COVID 19 (convalescent plasma). The procedure was uneventful but the patient’s condition became aggravated and he required orotracheal intubation and an increased requirement for vasoactive drugs. He also received broad spectrum antibiotics (linezolid and meropenem) and antifungal therapy (micafungin) but his situation worsened.

An echocardiogram showed severely impacted heart function. A bronchoalveolar lavage (BAL) identified a ventilator pneumonia due to *Klebsiella sp* and *Stenotrophomonas maltophilia*. Despite all these clinical complications his situation improved and he was transferred from the intensive care unit and discharged from the hospital.

A few days latter he developed a fever and a respiratory decline and was readmitted to the hospital. At this time his nasopharyngeal RT-PCR was still SARS-CoV-2 positive and, due to a respiratory decline raising concern of a fibrosing pneumonia, while on corticosteroids he also received intravenous immunoglobulin for one day accompanied by daily antibiotics. A BAL was negative. Voriconazole was prescribed due to a high galactomannan levels in the BAL, although no fungal infection was identified by culture. As his nasopharyngeal RT-PCR remained positive a second infusion of convalescent plasma was performed .This led to an increased lymphocyte count and clinical improvement and the patient was discharged from the hospital.

Due to the patient’s impaired immunological status, persistence of symptoms and prolonged positive RT-PCR result, it was decided to investigate the replicative capacity of his SARS-CoV-2 infection. From day 134 of illness until day 218 of illness, blood, urine, saliva, nasopharyngeal and anal swabs samples were collected at one-week intervals for analysis.

## Methods

From day 134 to day 218, blood, urine, saliva, nasopharyngeal and anal swabs were collected weekly. Swab samples from the nasopharynx, collected at the hospital from day 6 to day 120, were sent to the Virology Laboratory at Instituto de Medicina Tropical, São Paulo, Brazil. All samples collected were sent for viral identification, isolation and serological analyses. RNA extraction was performed using the QIAamp viral RNA kit, according to the manufacturer instructions. The RT-PCR assay was performed with the commercial RealStar® SARS-CoV-2 RT-PCR Kit 1.0, develop by *Altona Diagnostics*. DNA amplification employed the Roche LightCycler® 96 System.

### Virus isolation

For virus isolation, all clinical specimens that tested positive for SARS-Cov-2 by RT-PCR were inoculated into cultures of Vero cells (ATCC, CCL-81). Cells were seeded in a 25 cm^2^ cell culture flask in Dulbecco Minimal Essential Medium - fetal bovine serum and incubated overnight at 37°C. The next day, clinical specimens were added into the culture flask and incubated in a humidified 37°C incubator in an atmosphere of 5% CO_2._ The cultures were observed daily for 3-5 days for the presence of cytopathic effects (CPE). The supernatant was then collected, and virus replication was confirmed through CPE and by RT-PCR. Results of the SARS-CoV-2 isolation in Vero cells from clinical specimens are presented in Table 1.

**Table 1:** Weekly molecular detection of SARS-CoV-2 from different clinical specimens and samples (9 September 2020 to 9 April 2021).

| Date of sample collection | Clinical Specimen | PCR SARS-CoV-2 (Ct) of clinical specimen |  | GS | CPE after the 1st passage in culture | PCR SARS-CoV-2 (Ct) after 2<sup>nd</sup> passage in culture |  | Final result viral isolation (number of culture passages) |
| --- | --- | --- | --- | --- | --- | --- | --- | --- |
|  |  | Gene E | Gene S |  |  | Gene E | Gene S |  |
| Day 6 | NFS | 24.8 | 25.2 | T1 | Positive | 29.6 | 26.5 | Positive (2) |
| Day 77 | NFS | 21.1 | 21.1 | T2 | Positive | 26 | 28 | Positive (2) |
| Day 107 | NFS | 32.6 | 30 |  | Negative | ND | ND | Negative (2) |
| Day 128 | NFS | 31.5 | 31.7 |  | Positive | 35.93 | ND | Positive (2) |
| Day 134 | NFS Serum | 16.6<br>35.3 | 16.9<br>35.5 | T5 | Positive | 19.9<br>ND | ND<br>ND | Positive (2) |
| Day 141 | NFS Saliva Serum | 14.9<br>30.0<br>32.7 | 15.2<br>30<br>ND |  | Positive<br>Positive<br>Negative | 12.5<br>33<br>ND | 11.6<br>32<br>ND | Positive (2)<br>Positive (2)<br>Negative (2) |
| Day 148 | NFS Saliva Serum | 18.5<br>20.8<br>32.7 | 17.5<br>21.0<br>32.8 |  | Positive<br>Positive<br>Negative | 22<br>25<br>ND | 21<br>24<br>ND | Positive (2)<br>Positive (2)<br>Negative (2) |
| Day 155 | NFS Saliva Anal Swab Serum | 16.5<br>31.2<br>34.8<br>37.2 | 16.9<br>31.3<br>ND<br>38.7 |  | Positive<br>Positive<br>Negative<br>Negative | 16<br>34.9<br>ND<br>ND | 15<br>33<br>ND<br>ND | Positive (2)<br>Positive (2)<br>Negative (2)<br>Negative (2) |
| Day 162 | NFS Saliva Urine | 21.0<br>37.0<br>34.9 | 22.1<br>ND<br>ND |  | Positive<br>Negative<br>Negative | 23<br>ND<br>ND | 21.9<br>ND<br>ND | Positive (3)<br>Negative (2)<br>Negative (2) |
| Day 169 | NFS Saliva Urine | 15.7<br>31.0<br>34.7 | 15.6<br>30.7<br>ND | T10 | Positive<br>Positive<br>Positive | 19<br>ND<br>ND | 18<br>ND<br>ND | Positive (3)<br>Negative (2)<br>Negative (2) |
| Day 196 | NFS | 23.8 | 23.6 | T11 | Negative | ND | ND | Negative (2) |
| Day 218 | NFS | 35.1 | 36.4 |  | Negative | ND | ND | Negative (2) |

ND, Not Detected; NFS, Nasopharyngeal swabs; GS, Genome Sequencing; Ct, cycle threshold

### Virus Neutralization Test (VNT)

The presence of anti-SARS CoV-2 antibodies was determined using a cytopathic effect (CPE)-based virus neutralization assay. SARS-CoV-2 (EPI_ISL_1557222) was added to 96-well microtiter plates containing 5 × 10^4^ Vero cells/mL. This CPE VNT was adapted from Nurtop et al. 2018[4].

### SARS-COV-2 genomic sequencing and molecular analysis

The SARS-CoV-2 genomes from Day 6 (T1), Day 77 (T2), day 134 (T5), day 169 (T10) and day 196 (T11) were sequenced using multiplex PCR method as described previously [5,6]. Briefly, we used the open COVID-19 sequencing available and the bioinformatics protocols developed by the ARTIC network, described in detail at https://artic.network/ncov-2019. Reads were mapped against reference sequence Wuhan-Hu-1 (GenBank Accession Number MN908947) and low coverage regions were masked with N characters. Genomes were classified using the Pango lineage nomenclature system (http://pangolin.cog-uk.io/) [7] and a Maximum likelihood (ML) phylogenetic analysis was performed using complete reference genomes IQtree v2 [8] (Figure Supplementary). Consensus sequences generated in this study are available in GISAID under IDs EPI_ISL_1857098, EPI_ISL_1857094, EPI_ISL_1857095, EPI_ISL_1857096, EPI_ISL_1857097.

## Results

All NFS samples collected between day 6 and day 218 were positive for SARS-CoV-2 (a period of 218 days of continuous virus shedding). Samples collected after this date were negative for SARS-CoV-2 RNA. For saliva samples collected between day 134 and day 218, 5 of 8 were RNA negative. For serum samples collected in the same period, 4 of 8 were virus-positive. In addition, 2 of 8 urine samples and 1 of 8 anal swabs were virus-positive. Of the of 44 clinical samples sent for virus isolation, 12 were positive after their second passage: 9 from NFS and 3 from saliva (Table 1). Replicating virus was detected from NFS or saliva samples for up to 196 days.

Sera obtained on 10 occasions beginning on day 134 were consistently negative for neutralizing antibodies, suggesting acquisition of mutations that confer resistance to neutralizing antibodies (9).

Virus genomes from samples obtained on day 6 (T1), day 77 (T2), day 134 (T5), day 169 (T10) and day 196 (T11) were generated using a well-described multiplex PCR approach. Assembled viral genomes achieved between 75% and 98% genome coverage with a depth of at least 20x reads and were all classified as belonging to the B.1.128 lineage that emerged in Brazil [10]. The phylogenetic tree estimated with all B.1.1.28 genomes available until T11 (*n*=1,300) indicated that the patient’s virus sequences clustered monophyletically with maximum statistical support (phylogenetic bootstrap support = 100%), a scenario consistent with long-term persistent infection of SARS-CoV-2 in a single individual (Figure Supplementary).The closest available virus sequence was from São Paulo (GISAID ID: EPI_ISL_722007), consistent with local acquisition of the primary infection (patient reported no travel history). Samples had five cluster-defining mutations (three synonymous and two non-synonymous). One of these missense mutations had an impact on two proteins since it was located in an overlapping region coding for both nucleoprotein and ORF9b products. Between T2 and T5, eight additional mutations were acquired of which 7 were nonsynonymous (Figure 1). Most nonsynonymous mutations were retained until the last time point and were in genes coding for non-structural proteins within the ORF1ab, which corresponds to > 70% of the SARS-CoV-2 genome. Interestingly, the viral strains at all time points after T1 had a deletion corresponding to three amino acids (L141-; G142-; V143-; Δ141-143) in the spike protein gene at the N-terminal domain of the S1 subunit (NTD-S1), distal to the receptor-binding site (Figure 1). At the last time point (T11) an additional deletion was present at residue 144 (Y144-; Δ141-144) (Figure 1).

**Figure 1.**
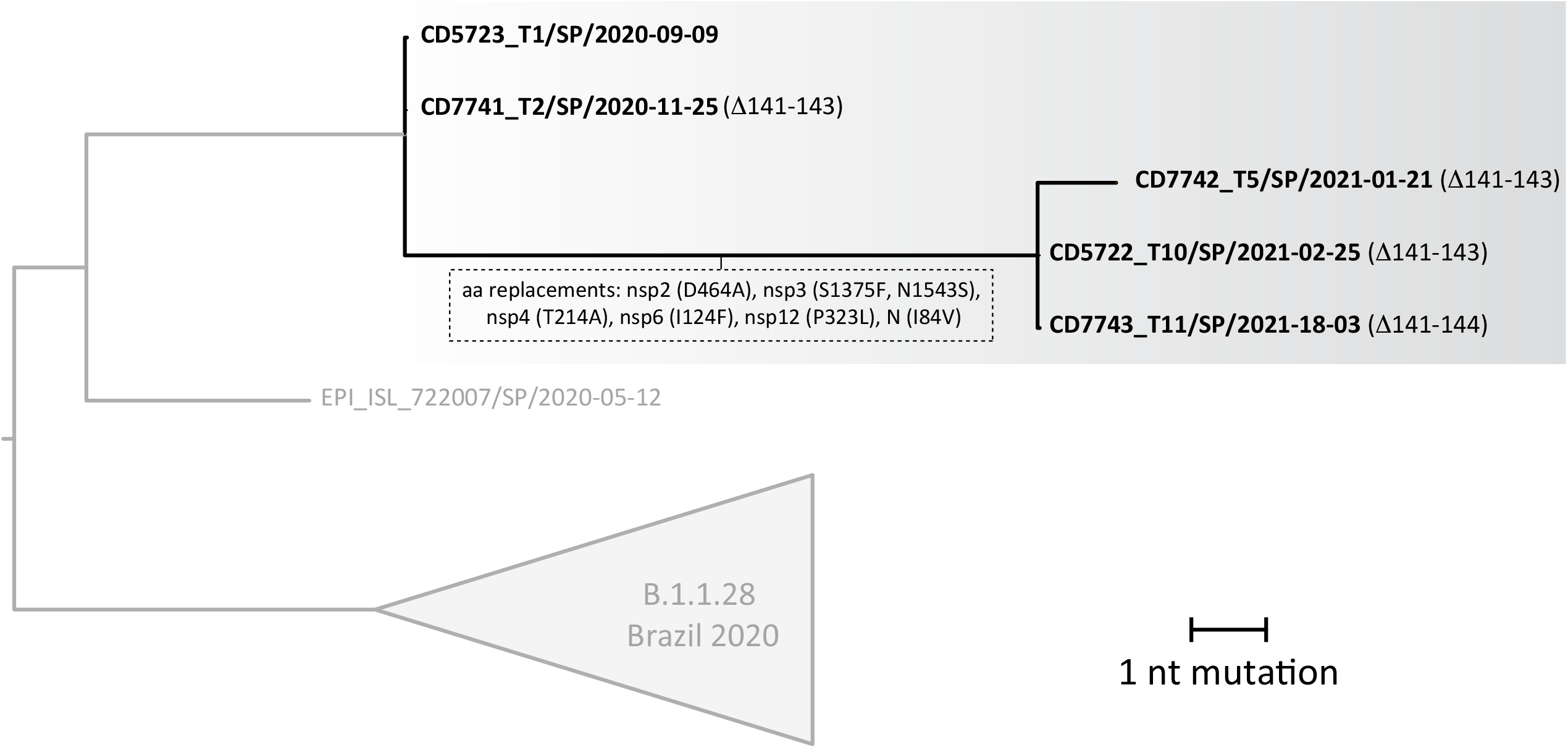
Zoom of the larger ML tree (Figure Supplementary) that includes all B.1.1.28 sequences from Brazil available in GISAID with date of collection <= T11

## Discussion

Here we report persistent SARS-CoV-2 shedding of infectious viruses in NFS and saliva samples collected for a period of over 196 days from an immunocompromised individual. Virus genome sequences collected >6 months apart confirm prolonged within-host evolution and reveal different mutations, including a convergent 4-amino-acid deletion (Δ141-144) in the N-terminal domain (NTD) of S1 (RDR2 region) that alters virus antigenicity [9].

Our phylogenetic analysis and long-term shedding of infectious virus confirmed through isolation in Vero cells, reveal an independent acquisition of a replication-competent, antigenically-distinct Δ141-144 variant. High frequency variants originating in individuals with prolonged shedding may spread and be selected at the population level, highlighting possible clinical and public health concerns regarding clinical management of immunocompromised individuals.

We detected several amino acid deletions within the spike protein. The Δ141-143 in-frame 3-amino-acid deletion in the N-terminal domain (NTD) was accrued after the first infusion of convalescent plasma, and between the first and second time point (between 7-22 days after the onset of the symptoms). This deletion was retained until last date of sample collection. At some point between T10 and T11, the virus acquired an additional adjacent deletion in the NTD, Δ144. Based on structural studies, the S1 and S2 subunits of the spike protein mediate receptor binding and membrane fusion and form the bulbous head and stalk region [11]. Although this region is supposed to be conformationally variable it is likely that changes at this region may impact protein binding to the cellular receptor [12]. Interestingly, deletions in NTD are not uncommon and have been repeatedly observed by others, in both immunocompromised individuals with prolonged shedding as well as in immunocompetent patients, and in viral strains belonging to distinct lineages [9,13]. The convergent NTD deletion observed here and elsewhere is highly suggestive of viral adaptation in response to convergent selective pressures. This is supported by the failure of virus neutralization by antibody 4A8 that targets the NTD region in *in vitro* tests [9].

To the best of our knowledge, this is the first case to report SARS-CoV-2 viral dynamics and viability in different clinical specimens for a prolonged period of time in an immunocompromised host. Studies to address the possible presence of SARS-CoV-2 in different clinical specimens in immunocompromised individuals, and utilization of culture techniques to identify replication-competent viruses, remain scarce.

Our findings may have implications for managing SARS-CoV-2 in long-term chronically infected individuals both in community and/or health care settings. Although SARS-CoV-2 RNA was detected in the majority of clinical specimens analyzed, including nasal secretions, saliva, serum, urine and anal samples, the importance of viral transmission from these last three sources seems to be at most marginally relevant to clinical practice. While we were only able to detect replication-competent viruses in nasal secretions and saliva, we cannot rule out that this may have been partially due to differences in viral titer in the various samples or to the presence of replication inhibitors. Nevertheless, our data suggests that considerations related to patient isolation should focus on viral presence in respiratory secretions and saliva in symptomatic patients.

A limitation of our study is that we were not able to test the contacts of the index person. Genome sequencing of viral isolates from clinical samples taken 196 days apart allowed us to observe changes in the SARS-CoV-2 genome over this prolonged time period. Studies investigating whether viral strains with the recurrent deletions associated with immune escape observed here can be effectively transmitted at the population level are urgently needed. Implementation of contact tracing, particularly for infections with longer generation intervals as described here, and continued monitorization of SARS-CoV-2 recent cases in Brazil will help to control community transmission of SARS-CoV-2 variants with altered epidemiological characteristics.

In conclusion, the present study demonstrates that viable and replication competent SARS-CoV-2 virus can be recovered from pharyngeal mucosa and saliva at prolonged intervals in an immunocompromised patient. The present observations may be pertinent to the further refinement of prevention and transmission protocols.

## Data Availability

The data that support the findings of this study are available from the corresponding author, upon request.

## Ethics

The study was approved by the local ethics committee :Comissao Nacional de Etica em Pesquisa do Ministerio da Saude do Brasil (CONEP), protocol No. 30419320.7.0000.0068, dated April 18, 2020) and all subjects provided informed written consent. Ethical approval was given to the project.

All necessary patient/participant consent has been obtained and the appropriate institutional forms have been archived.

## Acknowledgments and Funding

We are deeply grateful to the patient for allowing us to collect all samples, for his willingness and generosity to participate in the study and to provide written informed consent. We are grateful for the support of São Paulo Research Foundation (FAPESP) (Number:2020/05623-0). This work was also supported by a Medical Research Council-São Paulo Research Foundation (FAPESP) CADDE partnership award (MR/S0195/1 and FAPESP 18/14389-0) (caddecentre.org/). N.R. F is supported by Wellcome Trust and Royal Society (Sir Henry Dale Fellowship: 204311/Z/16/Z).

## Figure Supplementary-. Legend

**Figure Supplementary.**
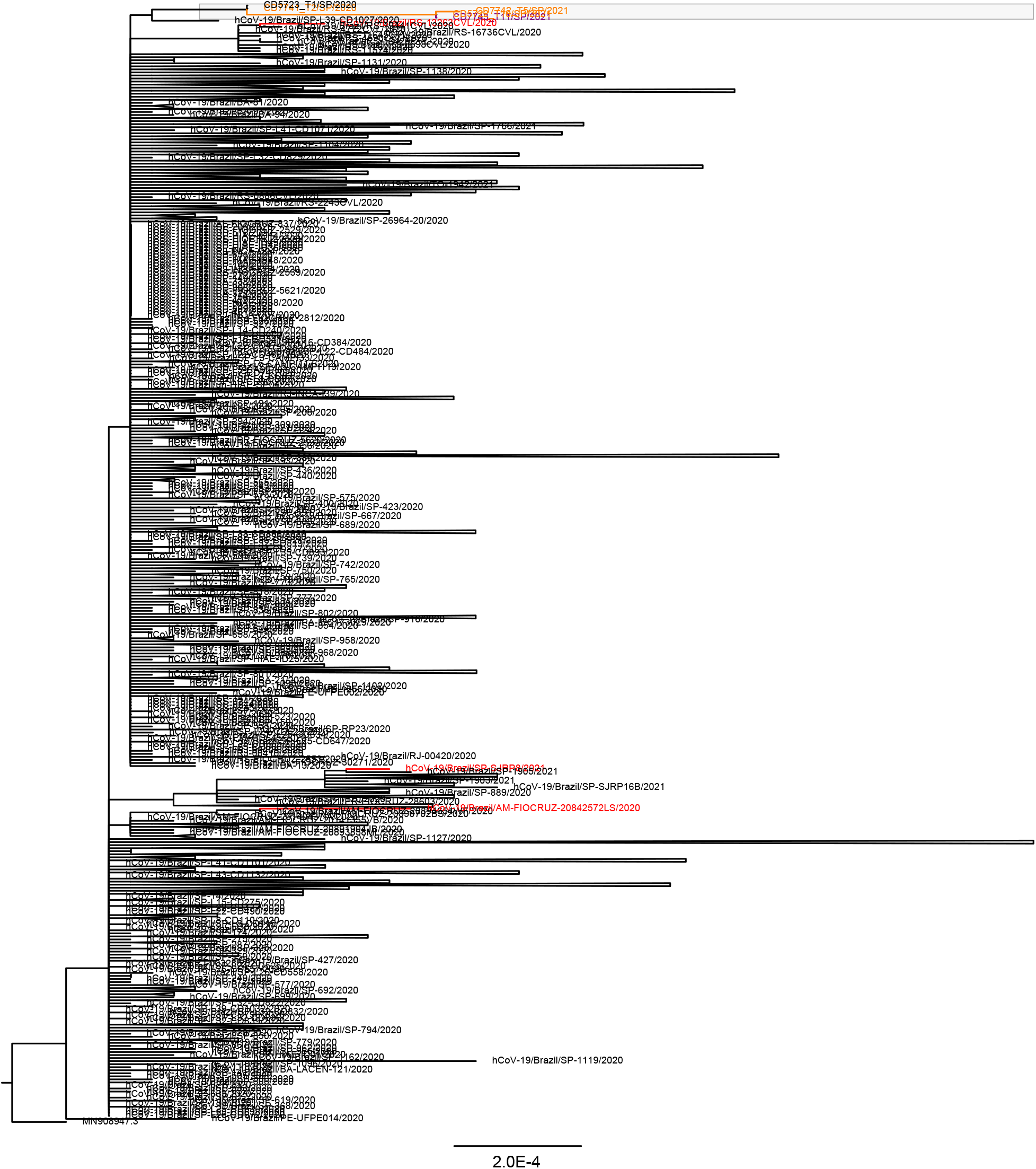
Full ML that includes all B.1.1.28 sequences from Brazil available in GISAID with date of collection <= T11. Red branches correspond to those with deletion 144 in spike, orange branches correspond to deletions 141-143 in spike, purple branches to deletions 141-144 in spike.

